# Infodemiological Study of Internet Search Pattern Related to Nipah Outbreaks in Bangladesh (Jan 2018- Jul 2023): A Google Trend Analysis

**DOI:** 10.1101/2025.07.02.25330742

**Authors:** Immamul Muntasir, M. Shafiqur Rahman

**Affiliations:** Institute of Epidemiology, Disease Control & Research (IEDCR), Mohakhali, Dhaka, Bangladesh; Institute of Statistical Research and Training (ISRT), University of Dhaka, Bangladesh

**Keywords:** Nipah virus, Outbreaks, Google trend, Bangladesh, Infodemiology, Internet

## Abstract

**Introduction:** Nipah virus is a fatal bat-borne pathogen that follows a seasonality from December to May in Bangladesh. Since 2001, Nipah outbreaks have been reported annually in Bangladesh. However, in early 2023, there has been an occurrence of a series of outbreaks here. This study aims to investigate the infodemiological aspect of this series of Nipah outbreaks by analyzing the Google search interest in Bangladesh.

**Methods:** The “Explore” feature of Google Trends was utilized to analyze search behavior focusing on the topics “Nipah virus infection” and “Date Juice” in Bangladesh from January 2018 to July 2023. Data from Nipah outbreaks during the same period was obtained. Correlation analysis was done between Relative Search Volume (RSV) and outbreak frequency, and spatial analysis to compare heat maps showing RSV and outbreaks.

**Results:** A line graph depicting the relative search volume (RSV) of Nipah virus infection reveals fluctuations in public interest, with spikes following outbreak events and during the Nipah season. Similarly, the RSV of “Date Juice” showcases changing patterns, occasionally aligning with Nipah outbreaks. Pearson correlation analysis indicates moderate positive correlations between Nipah-related RSVs and outbreaks, with p-values < 0.01, underscoring the link between public interest and outbreak frequency. Heat maps depict regional variations, with higher RSV regions coinciding with reported outbreaks.

**Conclusion:** The study found that RSV of both “Nipah Virus Infection” and “Date Juice” increased with the frequency of Nipah Outbreaks. We recommend continuous monitoring of health information regarding Nipah and other important public health issues.

## 1. Introduction

Nipah virus (NiV) is a pandemic potentially fatal zoonotic pathogen. NiV infection occurred when a spillover event happened from bat to human.^1–3^ For Bangladesh, the primary modes of NiV transmission involve: (i) consuming NiV-contaminated raw date palm sap (DPS) or date juice, (ii) human-to-human transmission, and (iii) exposure to sick animals.^4–7^ Here, NiV outbreaks follow a seasonal pattern, typically occurring between December and May. These outbreaks have been reported annually since 2001, coinciding with the harvesting season of date palm sap from November to March. Infections seem to be concentrated in the central and northwestern regions of Bangladesh, likely attributed to a higher prevalence of DPS consumption within this area.^8^

NiV was initially discovered as the cause of outbreaks of encephalitis cases in humans in Singapore and Malaysia from 1998 to 1999.^9^ Subsequently, it was identified as the agent behind encephalitis outbreaks in India and Bangladesh in 2001.^10,11^ Since then, NiV has continued to trigger human outbreaks in Bangladesh almost annually. A decrease in cases has been observed since 2016, attributed to extensive risk communication regarding the danger of drinking DPS. However, during January and February of 2023, 11 cases have been identified in two divisions of Bangladesh.^12,13^

Meanwhile, with the rise in worldwide internet usage, online searches have become a prevalent means of accessing health-related information.^14^ These searches offer not only a valuable tool for individuals seeking health insights but also hold significance for the scientific realm.^15^ This is because search queries can provide geographic and timely data about disease outbreaks, benefiting both individual users and the broader scientific community.

Infodemiology is the study of information regarding its distribution and influencing factors related to population or electronic media. Its ultimate goal is to bring information to the door of the policy level. One crucial aspect of infodemiology is to study online search patterns regarding health-related issues so that important insights can be derived for making decisions.^16^

Bangladesh has an increasing online presence, as seen by its internet penetration rate of 38.9% at the beginning of 2023. This represents an active internet-user base of 66.94 million people. Kepios data shows a notable increase of 691 thousand new internet users between 2022 and 2023. This growing use of the internet is impressive in terms of our study, which explores the dynamics of online search behaviors.^17^

**Figure 1:**
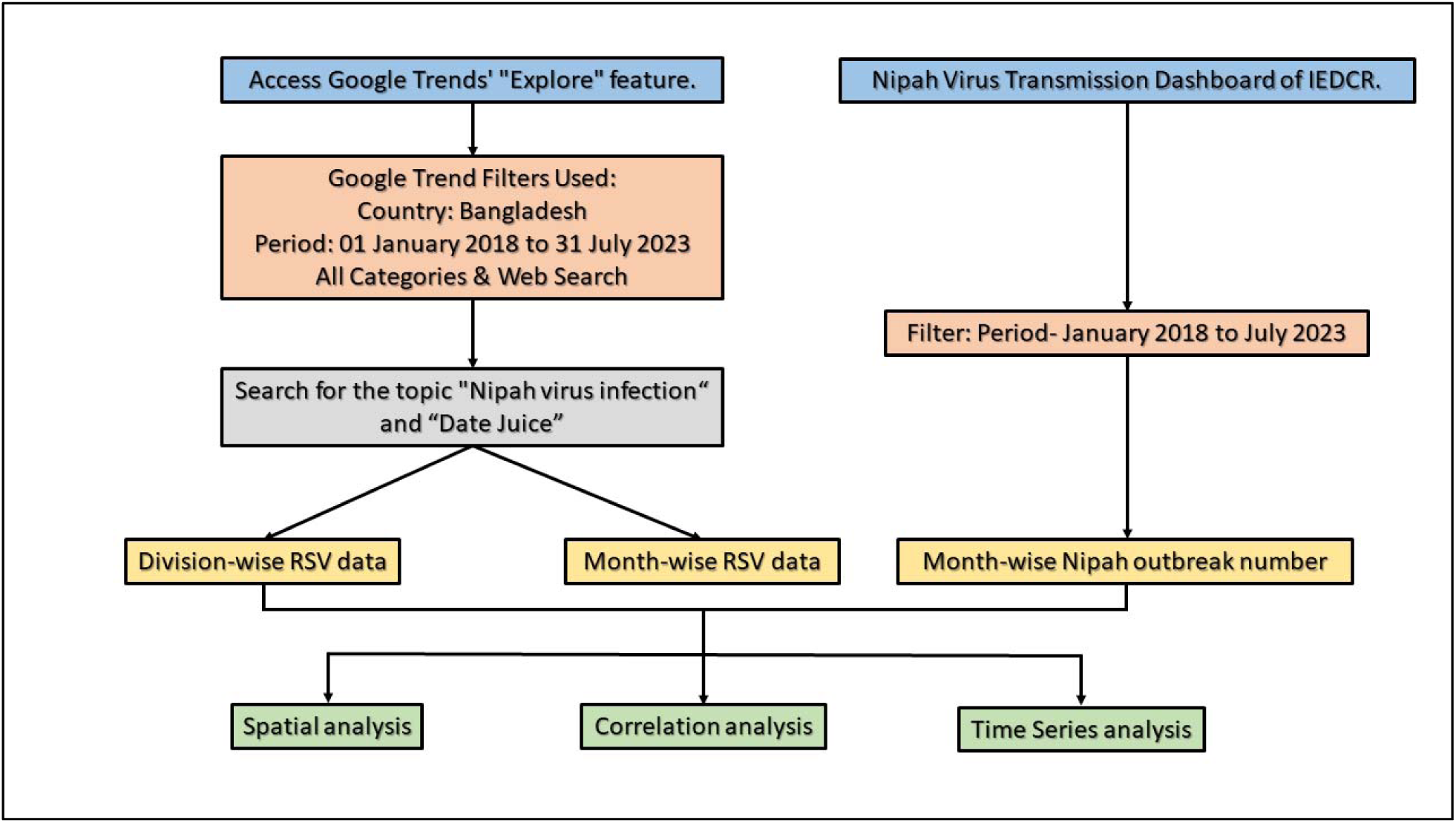
Methodology Flowchart for Analyzing Google Trends Data and Nipah Outbreaks

As the most common search engine is Google,^18^ the scientific community usually utilizes Google Trends to demonstrate and analyze the search interest of queries in various languages and places. Google Trends measures the level of online interest in a specific keyword, like “football,” and provides a normalized value of “minimum 0 and maximum 100” coined as relative search volume. This value is derived from the proportion of keyword-related queries compared to total web queries.^19^

Google Trends offers useful insights into community behaviors and health-related issues, notably with regard to infectious diseases, mental health, substance use, and general practices. A systematic review was carried out by Nuti et al in 2014, emphasized the expanding utilization of Google Trends by both clinical & public health research, particularly in monitoring infectious diseases, especially in nations with widespread internet access. Notably, Google Flu Trends, a model based on Google Trends, accurately forecast increases in influenza-like illness (ILI) cases before standard monitoring tools, although, with evident limitations. ^20,21^

This study’s objective is to utilize Google Trends to explore Google search interest in the context of Nipah outbreaks from January 2018 to July 2023.

## 2. Methods

Google Trends enables analysis with customization of the search term, period, and geographical area. Once a search term is entered and relevant time and location parameters are set, Google Trends generates visual representations showing how the search term’s volume compares to its peak popularity within the specified timeframe (a value of 100). The percentage of searches for a keyword in a place over a certain period is used to determine relative search volume (RSV). An RSV of 100 signifies the highest ratio of searches for a particular topic compared to total Google queries, while a zero value means that the search term’s queries were less than 1% of its peak RSV at the given time.

Searches were conducted on Google Trends on 5th July 2020. For keywords, the topic was chosen over search term. Because “a search term is a specific result that only includes the relative search volume for all keywords in the query in a specific language. On the contrary, a topic refers to a group of search terms which has the same concept, in every language”.^19^

Therefore, first the “Explore” feature of Google Trends was used and the popularity of the topic “Nipah virus infection” and “Date Juice” was searched. During this exploration the region was set to “Bangladesh”, the time frame from January 2018 to July 2023 in “All categories.” Then the data were retrieved in Comma Separated Value (CSV) format. Data were also obtained regarding Nipah outbreaks in Bangladesh timeline within the same time period from the Nipah Situation Dashboard of Institute of Epidemiology, Disease Control and Research (IEDCR). Then RSV data were plotted against Nipah outbreak data over the same period.

### 2.1. Statistical Analysis

Correlation analysis was performed among monthly RSV of “Nipah virus infection”, “Date juice” and frequency of Nipah outbreaks by month. Also,spatial analysis with Quantum Geographical Information System (QGIS) was done to compare between heat map of topics and Nipah outbreaks within preset time and location.

## 3. Results

### 3.1. Time Series Analysis

The line graph “Relative Search Volume of Nipah Virus Infection (Bangladesh) Over Time” shows fluctuations in the public interest and awareness of the Nipah virus over the given period. Initially, there are low search volumes from January to March 2018. A sharp spike appears in June 2018. From September 2018 to November 2018, there is a decline in search volume. Another spike occurs in January 2019, followed by fluctuating search volumes in the subsequent months. The period from January 2020 to June 2020 sees varying interest. The highest peak of interest has been observed during early 2023. Overall, the line graph demonstrates the fluctuating levels of interest in the Nipah virus over time.

**Figure 2:**
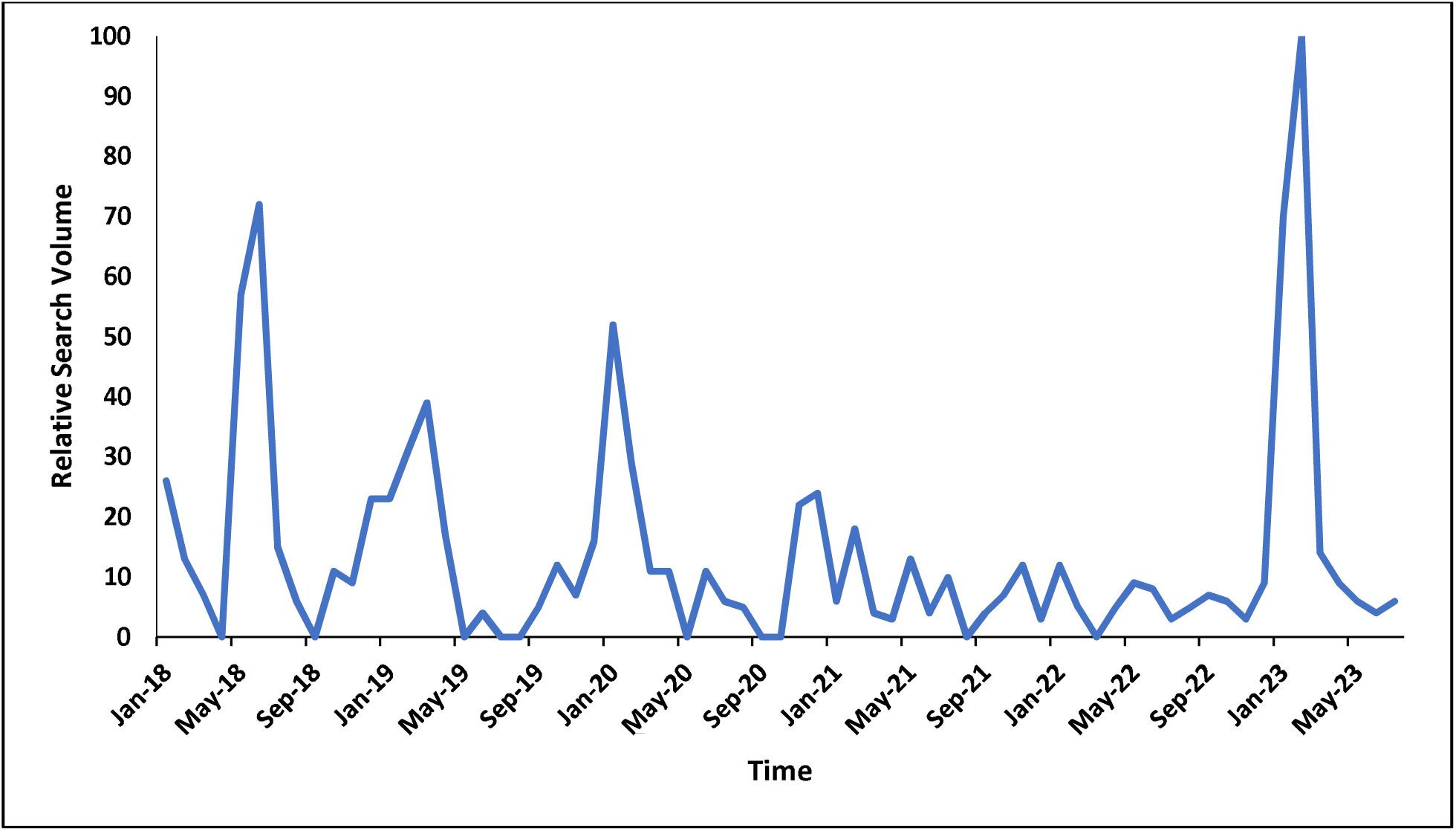
Relative Search Volume of Nipah Virus Infection in Bangladesh (Jan 2018 - Jul 2023)

**Figure 2:**
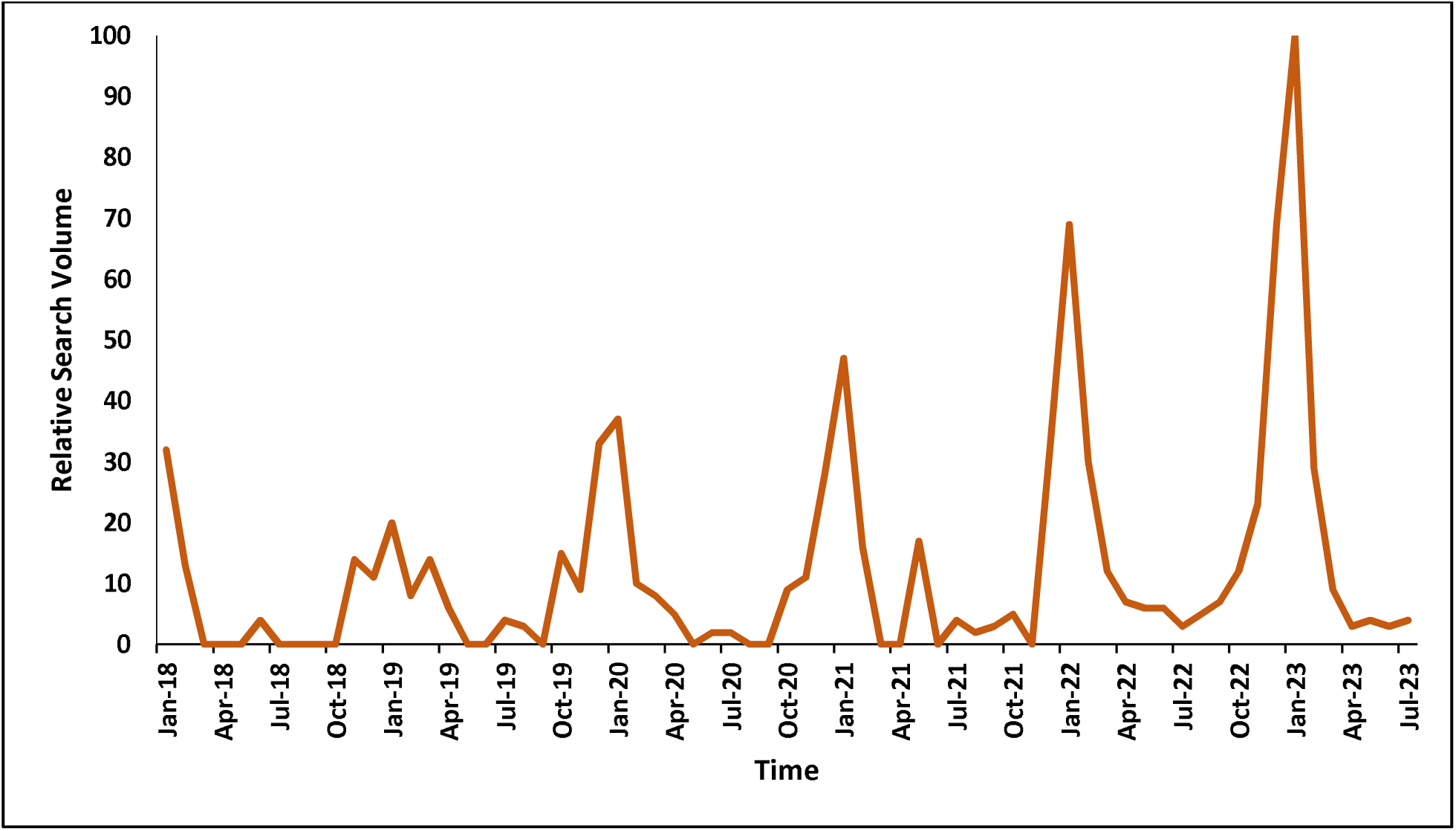
Relative Search Volume of “Date Juice” in Bangladesh (Jan 2018 - Jul 2023)

The line graph of “Relative search volume of date juice (Bangladesh) over time” illustrates a dynamic pattern of interest in date juice from Jan 2018 to Jul 2023.

The graph demonstrates that every year line graph of RSV of date juice starts to increase around the month of September or October and it reaches its peak during January, suggesting a seasonal pattern in interest regarding date juice consumption. The height of these peaks is also growing by the year, where the highest peak has been observed during early 2023.

**Figure 3:**
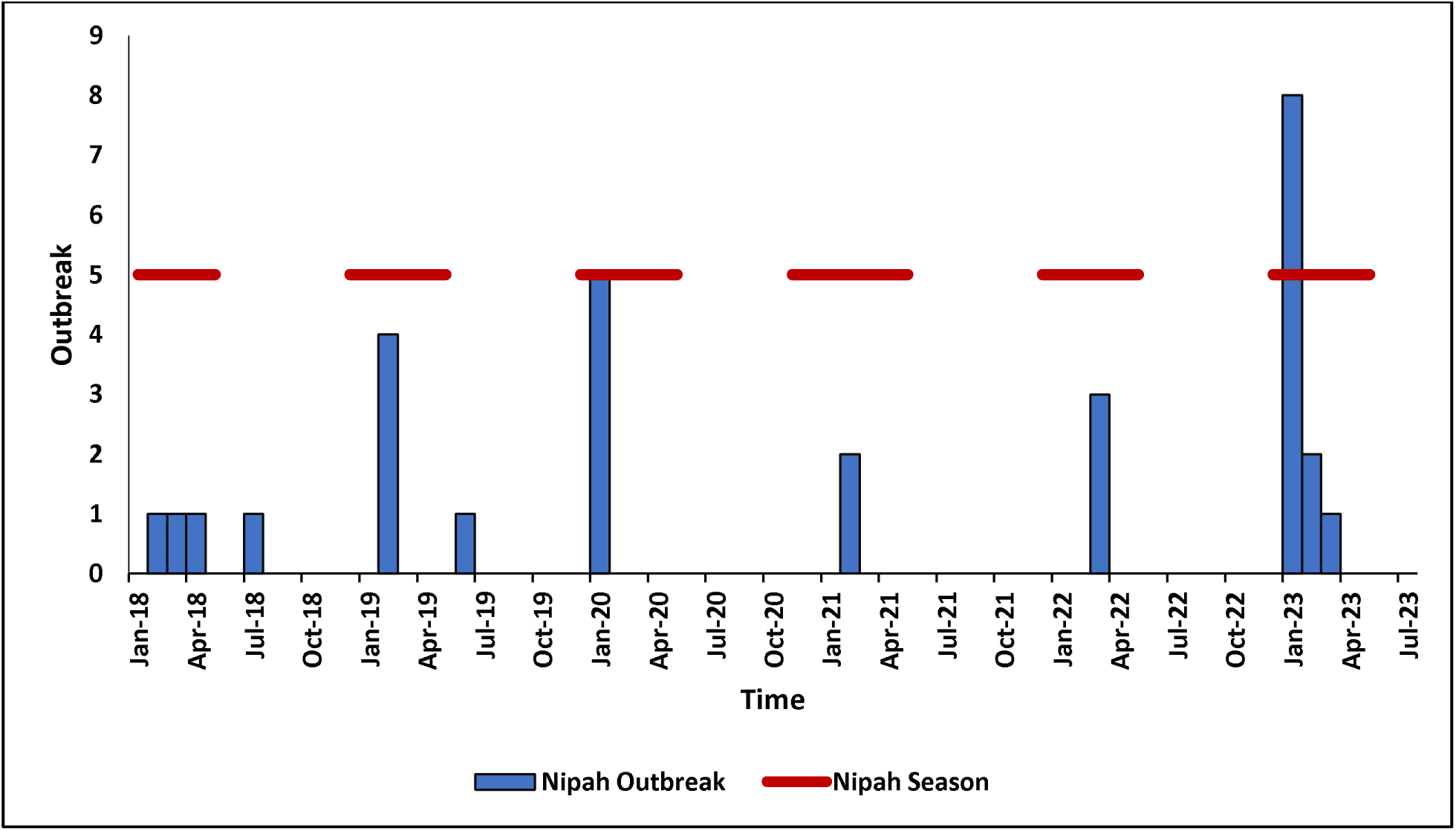
Nipah Virus Outbreaks in Bangladesh (Jan 2018-Jul 2023)

In this histogram, the vertical bars are representing the number of reported Nipah outbreaks in Bangladesh in each respective month which have been plotted against a timeline spanning from Jan 2018 to Jul 2023.

The chart demonstrates that during the Nipah season (December to May), there is a noticeable increase in outbreak frequency. Particularly, January 2020 and 2023 experienced the highest numbers of outbreaks. Other notable peaks have been observed in February 2019 and March 2022.

**Figure 5:**
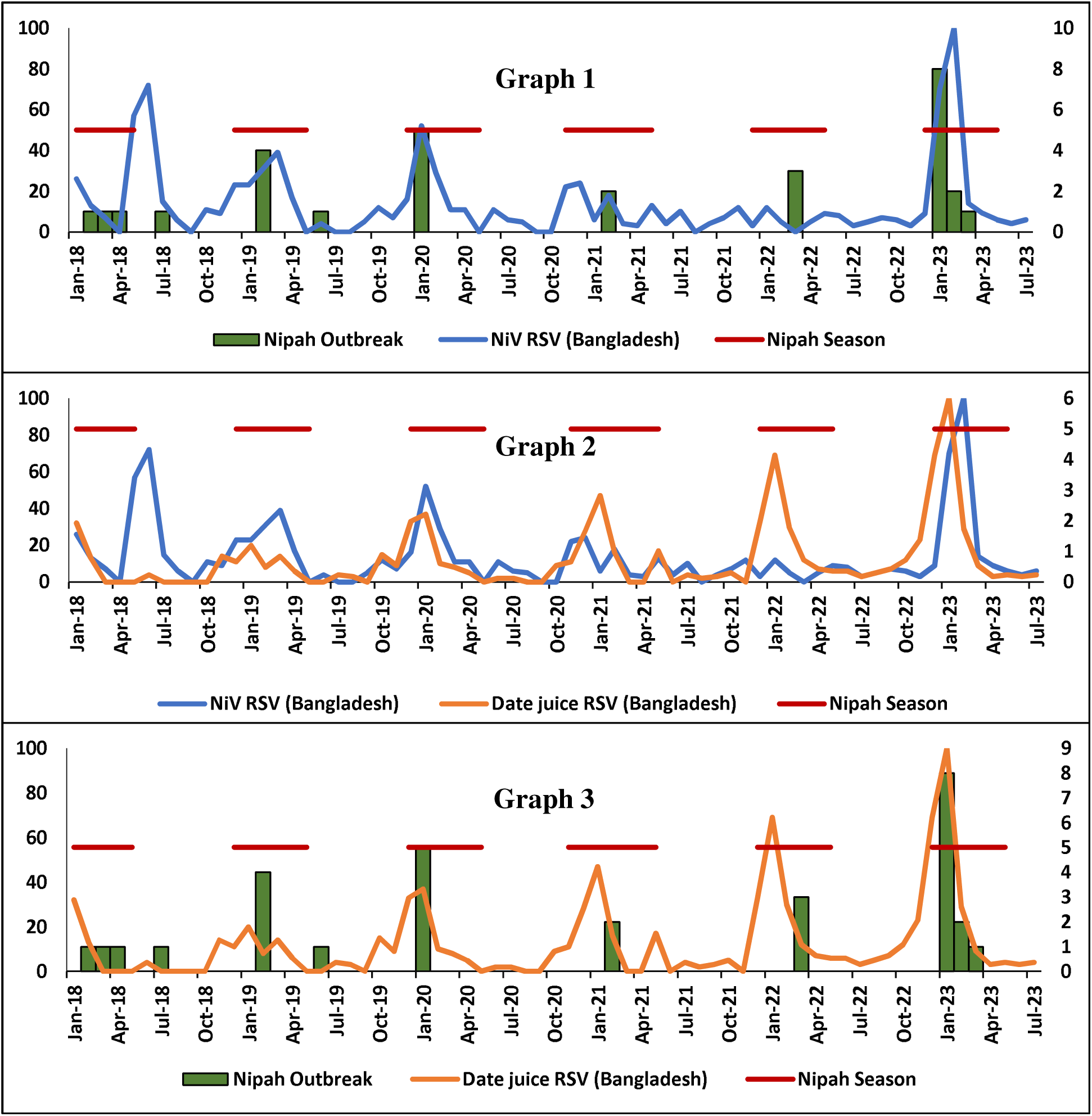
Relation Between RSV of “Nipah Virus Infection” and “Date Juice” and Nipah Outbreaks & Its Season in Bangladesh (Jan 2018-Jul 2023)

Here, the first graph illustrates the comparison among RSV, outbreaks, and seasonality of the Nipah virus. The graph implies that RSV tends to rise around the occurrence of an outbreak and during the Nipah Season (December to May).

The second graph shows the relation between the RSV of date juice and Nipah outbreaks and season. The graph also features several spikes in RSV during Nipah season and the notable ones happen to be around the time of multiple outbreaks. And the most prominent peak coincides with the series of outbreaks during early 2023.

Finally, the third graph compares the line graph of RSV of Nipah virus and date juice. Both line graphs illustrate some peaks in RSV around the same period. The time of the most prominent shared peak also coincides with the time of occurrence of the series of outbreaks of 2023.

### 3.2. Correlation Analysis

The Pearson correlation coefficient was calculated to explore the relationship between “RSV of Nipah Virus Infection,” “RSV of Date Juice,” and “Nipah Outbreak.” The calculated correlation coefficients and their significance levels (p-values) are presented in the table.

The calculated Pearson’s Correlation Coefficient values indicate a significant moderately positive correlation between the RSV of Nipah virus infection and date juice and the frequency of Nipah outbreaks.

**Table 1:**
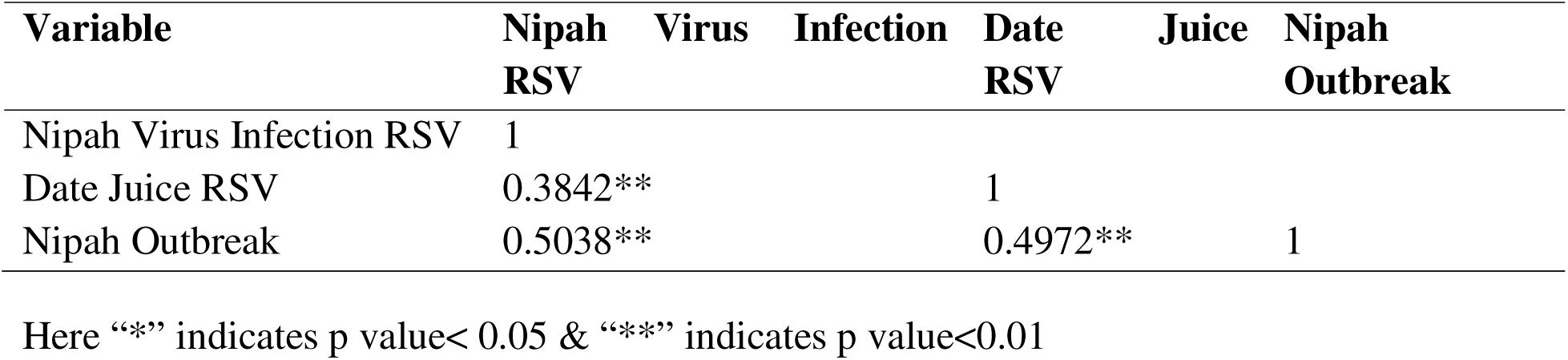
Pearson Correlation Coefficient Scores with P value of observed RSV and outbreaks related to Nipah Virus in Bangladesh (Jan 2018-Jul 2023)

### 3.3. Spatial Analysis

QGIS was used to plot spatial data regarding the RSV of Nipah virus infection and date juice and the number of outbreaks in different divisions of Bangladesh from January 2018 to August 15, 2023, and generated three heat maps.

The first map (Map-1) displayed that the divisions of Rajshahi and Dhaka stand out with high RSV values of 100 and 97 respectively, portraying significant public interest and awareness of Nipah virus infection in these regions.

Similarly, the second map (Map-1) showed that the division of Rajshahi had the highest number of outbreaks with 13 cases, followed closely by the Dhaka division with 12 outbreaks. The barisal division had 2 outbreaks, while the other divisions (Sylhet, Khulna, Chittagong, Rangpur) show no reported outbreaks. This heat map indicates a concentration of outbreaks in the Rajshahi and Dhaka divisions.

When these two heat maps were compared, it was observed that divisions with a high number of outbreaks had the highest RSV values for Nipah virus infection. Rajshahi and Dhaka divisions show prominent RSV values, they also have a notable number of outbreaks.

The third heat map (Map-2) illustrated the RSV values regarding “date juice” which indicated the divisions with highest RSV were Rajshahi, Dhaka and Khulna. When comparing with the previous two heat maps, the spatial pattern for RSV of “ date juice” in Dhaka and Rajshahi aligned with those of RSV for Nipah virus infection and number of outbreaks. However, a contrast was observed regarding Khulna division, where the highest RSV was observed in terms of date juice though its RSV for Nipah virus infection and number of outbreaks were low.

**Map 1:**
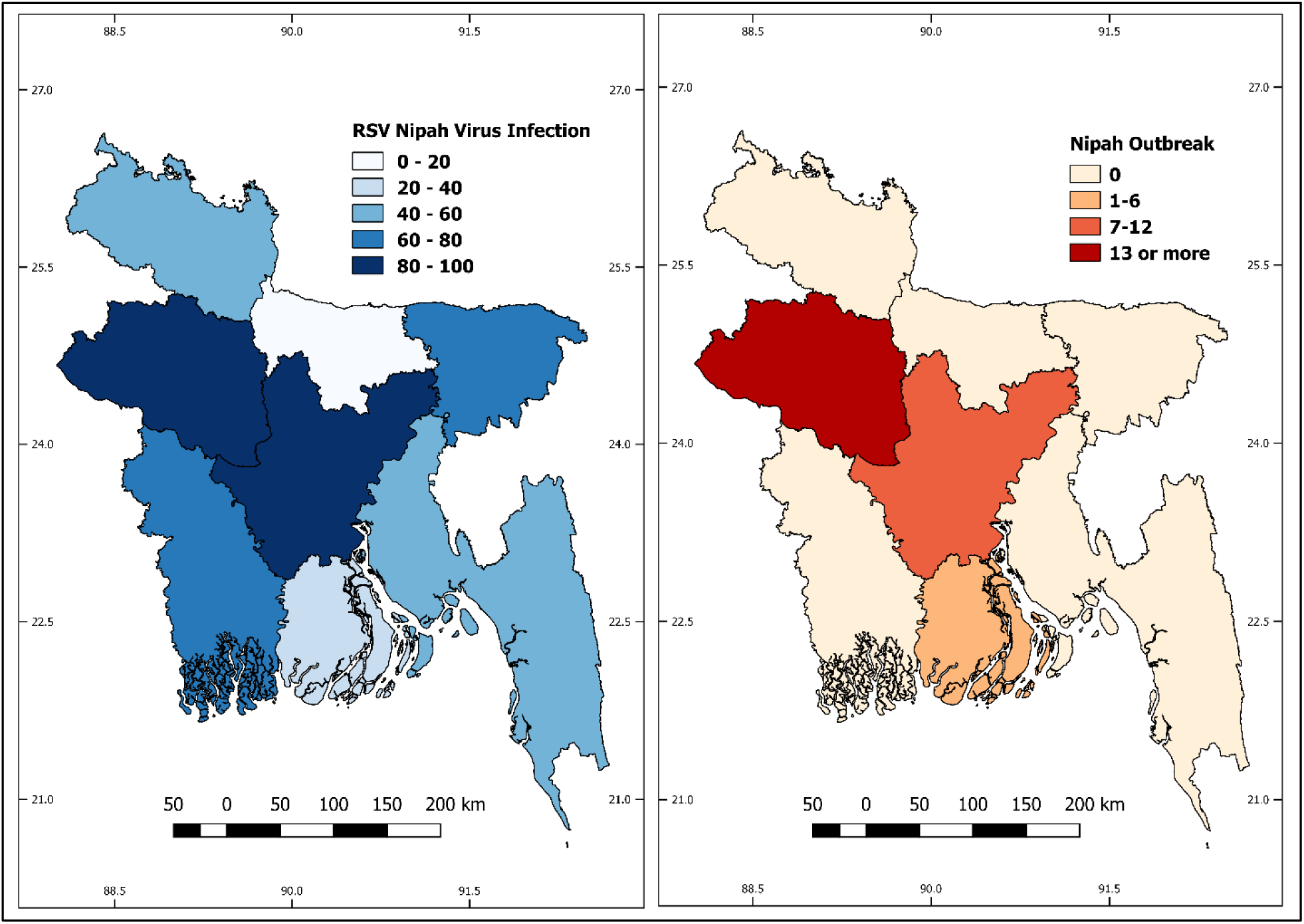
Maps Showing Distribution of RSV and Frequency of Nipah Outbreaks by Divisions of Bangladesh from Jan 2018 to Jul 2023

**Map 2:**
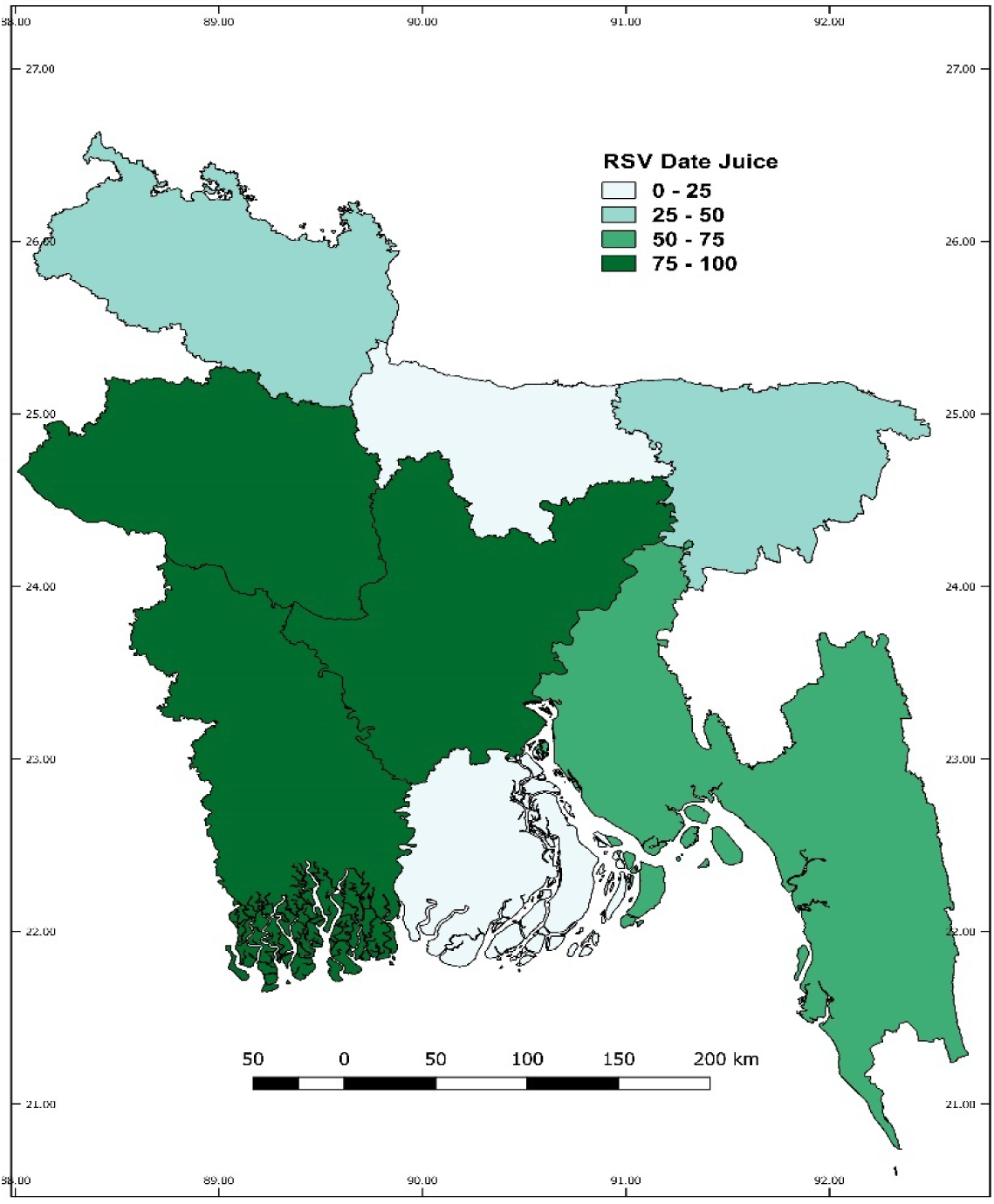
Maps Showing Distribution of Date Juice by Divisions of Bangladesh from Jan 2018 to Jul 2023

## 4. Discussion

It is evident now that Bangladesh is in constant threat of the Nipah virus. Therefore, our study focuses on people’s information-seeking behavior and pattern on the internet and how it links to Nipah outbreaks and seasonal trends. Our study revealed that people have been frequently searching for information about the Nipah virus in Bangladesh for over five and a half years.

During the middle of 2018, we noticed spikes. January 2019 saw a big jump, followed by ups and downs, showing changing interest over time. However, the most significant moment came in January 2023, when the graph hit an all-time high. This suggests recent intense interest, possibly due to new developments or media coverage.

Comparison of these findings to Nipah outbreaks and the “Nipah season,” suggested an connection. The period from December to May, known as “Nipah season,” sees more outbreaks. Interestingly, the graph’s ups and downs mirror the timing of these Nipah seasons. And also, the highest peak of interest for Nipah virus infection coincides with the record number of series of Nipah outbreak occurrences in the country. This was likely driven by healthcare professionals and public seeking information post-confirmation.

Another interesting aspect is observed regarding people’s searching for “Date Juice” in Bangladesh during the same time frame. The line graph in Figure-2 tells a story of changing interest in this topic. There was a cyclical peak during the winter season. These peaks were growing by year and the maximum peak of interest happened at the same time as the series of outbreaks in 2023. ^12^ However, we cannot derive any conclusion whether the “date juice” searching behavior can be a predictor or just an act of curiosity of mass people in the face of the outbreaks as the seasonal peaks reflect interest in commerce rather than virus transmission concerns.This along with correlation analysis suggests the RSV correlates with heightened awareness following confirmed cases.

Bringing these narratives together, a relationship between the line graphs, Nipah outbreaks, and Nipah seasonality, appeared. The ups and downs in the Nipah virus search volume align with the timing of outbreaks, indicating a proactive approach to seeking information during risky periods. Similarly, the shifts in interest in date juice, as seen in the second graph, respond to specific events, showing the interplay of curiosity and external factors.

Finally, the spatial analysis though reflects an alignment between RSV of nipah virus infection and number of Nipah outbreaks, it cannot be denied that regions with lower RSV may still have undetected or underreported cases.

As a zoonotic disease, Nipah prevention and control strategy requires a One Health approach through coordinated efforts across human, animal, and environmental health sectors, with robust surveillance to detect infections early in humans and animals. Biosecurity measures targeting the animal-human interface and public education to reduce exposure, such as avoiding raw date palm sap are cruical. Infodemiology complements this approach by monitoring online platforms and social media to detect misinformation, emerging concerns, and knowledge gaps about Nipah transmission and prevention. This real-time data enables public health authorities to design communication strategies, address misconceptions, and disseminate accurate information and ultimately enhance public adherence to preventive measures. ^22–25^

Therefore, these findings support the idea of monitoring public search behaviors and using social media data to understand evolving trends. The correlation between these line graphs and Nipah outbreaks and seasonal patterns highlights people’s tendency to seek information during times of higher risk. This has important implications for public health communication, emphasizing the need for timely and accurate information to counter misinformation. By uncovering these complex relationships, our study contributes valuable insights to addressing outbreaks and their impact on public health and well-being.

## 5. Limitations

Our study has some limitations. We only used Google to retrieve data. Therefore, there is a strong chance of missing information from other search engines. Additionally, our analysis was done on English search results. Finally, we didn’t have any access to demographic information such as age, sex, and education status of users, which eventually restricted our ability to fully comprehend the population.

## 6. Conclusion

Our study found that RSV of both “Nipah Virus Infection” and “Date Juice” increased with the frequency of Nipah Outbreaks. People actively seek out information when there is an imminent risk. We recommend continuous monitoring of health information regarding Nipah and other important public health issues and also, reinforcement of these monitoring during public health emergencies.

## 7. Author’s Contribution

Immamul Muntasir was responsible for the conceptualization and research design. Data collection and analysis were performed by all Authors. The initial manuscript draft was carried out by Immamul Muntasir. M. Shafiqur Rahman contributed to thorough review and editing of the final manuscript. All authors participated in final approval and made substantial contributions to the manuscript’s publication.

## Data Availability

All data produced in the present study are available upon reasonable request to the authors

## Acknowledgments

The authors are thankful to the Nipah Virus Transmission Dashboard of the Institute of Epidemiology, Disease Control & Research (IEDCR) under the Ministry of Health & Family Welfare (MOHFW).

